# Dosimetric comparison of IMPT versus IMRT in unilateral treatment of head and neck cancer

**DOI:** 10.1101/2025.09.23.25336270

**Authors:** Stephanie Zhao, Kevin Chen, Michael Watts, Kenneth Walker, Jessica Hilliard, Yao Hao, Stephanie Perkins, Anthony J. Apicelli, Nikhil Rammohan

## Abstract

**Purpose:** There is currently no consensus on the role of proton therapy in head and neck cancers. We conducted a retrospective dosimetric comparison of delivered photon-based intensity modulated radiation therapy (IMRT) plans with simulated intensity-modulated proton therapy (IMPT) plans.

**Patients and Methods:** In this single-institution retrospective review, we included patients with primary tumors from all head and neck sites treated with unilateral IMRT, who experienced worsened dysphagia and xerostomia symptoms post-radiation. MD Anderson Dysphagia Inventory (MDADI) and Xerostomia Questionnaire (XQ) scores were prospectively collected. We compared target coverage (V95%) for high-dose and low-dose clinical target volumes (CTVs) and maximum/mean doses for organs-at-risk (OARs) between delivered IMRT plans and simulated IMPT plans. Statistical analysis was performed using Wilcoxon signed-rank tests, with Bonferroni-corrected significance level of 0.003.

**Results:** A total of 23 patients were included in the study. Both IMRT and IMPT plans provided appropriate target coverage of the high-dose CTV (median V95 99.91% for both) and low-dose CTV (median V95 99.71% and 99.90%, respectively). IMPT plans allowed for significant reduction in maximum dose to critical OARs, including the spinal cord (6.4Gy vs 37.3Gy IMRT, p<0.001) and brainstem (5.6Gy vs 33.0Gy IMRT, p<0.001). Furthermore, mean dose to the oral cavity and contralateral pharyngeal constrictors were significantly reduced in IMPT plans (19.7Gy vs 33.6Gy IMRT oral cavity, p<0.001; 20.4Gy vs 26.2Gy IMRT contralateral pharyngeal constrictor, p<0.001). IMPT spared dose to the contralateral parotid (0.04Gy vs 7.6Gy IMRT, p<0.001) and contralateral submandibular gland (1.4Gy vs 15.4Gy, p<0.001).

**Conclusion:** IMPT spares dose to OARs compared to IMRT plans in head and neck cancers treated with unilateral radiation. We hypothesize that IMPT can reduce acute and long-term toxicity for these patients, even in locally advanced cancers. Future prospective comparison between these treatment modalities is indicated.

## INTRODUCTION

Intensity-modulated radiation therapy (IMRT) remains a cornerstone of treatment for head and neck cancers. However, toxicities from IMRT and other photon techniques are common, ranging from acute toxicities of mucositis, xerostomia, and dysphagia to long-term toxicities of osteoradionecrosis and permanent PEG-tube dependence [1]. In particular, late and potentially irreversible dysphagia and xerostomia have been shown to have a detrimental impact on quality of life in head and neck cancer survivors [2].

While photon-based linear accelerators remain the primary modality of modern radiation therapy, proton radiotherapy has emerged as an alternative approach with unique dosimetric characteristics. Intensity-modulated proton therapy (IMPT) utilizes the Bragg peak of protons to precisely deposit energy at the depth of the target and eliminate exit dose, thereby sparing healthy tissue and reducing dose to organs at risk (OARs) while maintaining target coverage. The potential benefit of IMPT is particularly pronounced for patients with lateralized disease treated with unilateral elective nodal irradiation, where contralateral OARs can potentially be spared more effectively. Prior dosimetric analyses of unilateral IMPT compared to IMRT demonstrated significantly reduced dose to OARs, including 75-90% reduction in mean dose to the oral cavity and >90% reduction in dose to the contralateral parotid and submandibular glands [3-6]. Furthermore, systematic reviews have confirmed the dosimetric advantages of IMPT across multiple head and neck sites, in patients treated with both bilateral [7] and unilateral irradiation [6].

Currently, there is limited evidence supporting IMPT in head and neck cancers requiring unilateral neck radiation. A recent Phase III trial comparing IMPT and IMRT in oropharyngeal cancer established the non-inferiority of IMPT in rates of progression-free survival at three years. Furthermore, there were lower rates of PEG-tube dependence and significant weight loss in the IMPT group compared to the IMRT group [8]. Other investigations have focused on reirradiation [9-12] or a single subsite [13, 14]. Despite evidence in favor of IMPT for head and neck cancer, there is concern that dosimetric advantages and patient-reported toxicity reduction of unilateral IMPT observed in the small number of prior studies were limited to those with less extensive disease burden [5]. That is, the extent of dosimetric benefit may not be as pronounced in cases which remain well-lateralized but are locally advanced, thus preventing any clinically meaningful dose sparing to nearby OARs.

In this study, we predominately included patients with locally advanced head and neck cancer across multiple subsites, and who experienced significant dysphagia and xerostomia after unilateral irradiation with IMRT. We then performed an *in silico* comparison to simulated IMPT treatment plans to determine if proton therapy was dosimetrically superior in this cohort characterized by more advanced disease and increased toxicity burden.

## METHODS

### Patient selection

In this retrospective chart review, we used a registry of head and neck cancer patients treated at our institution from 2006-2020 with prospectively collected, validated patient-reported outcome measures via the MD Anderson Dysphagia Inventory (MDADI) [15] and Xerostomia Questionnaire (XQ) [16]. Composite MDADI scores range from 20 (low functioning) to 100 (high functioning). XQ scores range from 0 to 100, and higher scores reflect more severe xerostomia. Patients with primary tumors from all head and neck sites, including cutaneous malignancies, treated with unilateral radiation with a ≥10 point decrease in MDADI composite score from pre- and post-radiation, or <60 point score on MDADI post-radiation, were included in the study. The MDADI thresholds for inclusion were chosen based on previous studies establishing a 10 point change in MDADI as clinically significant [17], as well as MDADI <60 as indicative of moderate/severe dysphagia [18]. All patients were treated with IMRT, either in the post-operative or definitive setting. Chemotherapy was used at the discretion of the treating medical oncologist, either as concurrent therapy with definitive radiation or adjuvant therapy for select patients with positive surgical margins and/or extranodal extension. Patients were included regardless of radiation intent (definitive vs post-operative) and receipt of chemotherapy. This study was approved by the institutional review board of the institution (IRB ID 202401089).

### Treatment planning and delivery

Computed tomography (CT) simulation was performed using a thermoplastic immobilization mask and arm stretchers. Image registration to diagnostic imaging was done when available to aid in target delineation. The primary tumor/tumor bed and involved lymph nodes were contoured as the gross tumor volume (GTV). A high-risk clinical target volume (CTV) was generated by applying a 1 cm expansion to the GTV while respecting anatomical boundaries. An elective nodal volume was delineated following consensus guidelines. A planning target volume (PTV) was generated by applying an isotropic 5 mm expansion to the CTV. Radiation doses were prescribed to the PTV. For definitive radiation, 70Gy was prescribed to the high-risk target volume, with 56Gy to the elective nodal volume, via a simultaneous integrated boost (SIB) technique. For post-operative radiation, 60-66Gy was prescribed to the high-risk volume and 52-54Gy to the elective nodal volume.

Conventional x-ray radiotherapy was delivered using a linear accelerator (LINAC) on the Varian TrueBeam to generate 6 or 10 MV photon beams. Volumetric modulated arc therapy (VMAT) treatment plans were generated via inverse planning in Eclipse.

### IMPT plan generation

Proton-based planning employed robust optimization of pencil beam scanning, which is delivered by the Mevion S250i HYPERSCAN proton machine. Treatment plans were generated within Raystation version 12A with full Monte Carlo dose calculation (Version 5.4). Multi-field optimization was conducted directly on the CTVs with applied range uncertainty of 3% and isotropic setup uncertainty of 5 mm for all cases. Three-field proton plans with static aperture technique were utilized with target coverage as the primary factor in plan comparison with photon-based plans.

### Dosimetric comparison and statistical analysis

V95(%) for high-dose and low-dose CTVs were reported for each IMRT (treated) and IMPT (simulated) plan. OARs associated with common treatment toxicities were included, namely the esophagus, larynx, oral cavity, lips, bilateral pharyngeal constrictors, bilateral parotid glands, and bilateral submandibular glands. The brainstem and spinal cord were also included as relevant central nervous system structures. For serial critical structures, such as the spinal cord and brainstem, the maximum doses were reported for the IMRT and IMPT plans. The maximum dose was also reported for a 3 mm skin rind to investigate differences in skin dose between IMRT and IMPT. For parallel OARs, mean doses were compared.

We assessed differences in pre- and post-radiation therapy (RT) scores on the patient-reported outcome measures of MDADI and XQ. Given the nature of the study’s inclusion criteria, certain patients did not have paired MDADI scores. For those patients, their pre-RT MDADI scores were imputed with the median of all pre-RT MDADI scores. The same process was taken for patients without pre- and post-RT XQ scores. Scores for MDADI and XQ pre- and post-RT were compared with a Wilcoxon signed-rank test. For dosimetric comparisons of V95 and OARs, we reported median of the mean/max doses for each radiation plan and evaluated differences using a Wilcoxon signed-rank test. We employed a Bonferroni correction for multiple comparisons, with an adjusted significance level of 0.003.

## RESULTS

Twenty-three head and neck cancer patients treated with unilateral IMRT who experienced worsened dysphagia symptoms post-radiation were included in the study. Characteristics of patient demographics, tumors, and treatment plans are presented in **Table 1**. Ages of the patients ranged from 37 to 86 years. Fifteen patients (65%) had primary tumors in the oropharynx, while other less common subsites included oral cavity (n=1), parotid gland (n=2), and cutaneous malignancies (n=2). Over half of the patients in the cohort (57%) had locally advanced disease with either T3-T4 or N2-N3 disease. Most patients received post-operative radiation compared to definitive radiation (91% vs 9%, respectively). The median pre-RT MDADI score was 81 (range 48-98), and the median post-RT MDADI was 56 (range 37 to 76), which was significantly different (p<0.001). Similarly, XQ scores were significantly different (p<0.001), with higher post-RT scores reflecting worsened xerostomia following radiation.

**Table 1.** Patient characteristics.

| **Characteristics** |  |
| --- | --- |
| Age at diagnosis, years, median (range) | 60 (37-86) |
| Sex, n (%) |  |
| Male | 19 (83%) |
| Female | 4 (17%) |
| Race |  |
| Caucasian | 21 (91%) |
| Black/African American | 2 (9%) |
| Smoking History, n (%) |  |
| Yes | 15 (65%) |
| No | 8 (35%) |
| Disease Site |  |
| Oropharynx | 15 (65%) |
| Oral Cavity | 1 (4%) |
| Parotid | 2 (9%) |
| Skin | 2 (9%) |
| Other/Unknown Primary | 3 (13%) |
| T stage<sup>a</sup>, n (%) |  |
| 1 | 5 (22%) |
| 2 | 8 (35%) |
| 3 | 3 (13%) |
| 4 | 2 (9%) |
| TX/T0 | 5 (22%) |
| N stage, n (%) |  |
| 0 | 2 (9%) |
| 1 | 8 (35%) |
| 2 | 9 (39%) |
| 3 | 1 (4%) |
| NX | 3 (22%) |
| Received chemotherapy, n (%) |  |
| Yes | 7 (30%) |
| No | 16 (70%) |
| Radiation Therapy |  |
| Dose |  |
| < 60 Gy | 2 (9%) |
| 60 Gy | 10 (43%) |
| 66 Gy | 9 (39%) |
| > 66 Gy | 2 (9%) |
| Intent |  |
| Post-operative | 21 (91%) |
| Definitive | 2 (9%) |
| Quality of Life |  |
| MDADI<sup>b</sup> |  |
| Pre-RT score, median (range) | 81 (48-98) |
| Post-RT score, median (range) | 56 (37-76) |
| Xerostomia questionnaire<sup>c</sup> |  |
| Pre-RT score, median (range) | 11.5 (0-33) |
| Post-RT score, median (range) | 47 (8-76) |
| Required G-tube, n (%) |  |
| Yes | 5 (22%) |
| No | 18 (78%) |

a. AJCC 8<sup>th</sup> edition
b. MD Anderson Dysphagia Inventory, scores range from 20 (worst function) to 100 (best function).
c. Xerostomia questionnaire, scores range from 0 (no xerostomia) to 100 (severe xerostomia)

The target coverage and OAR doses are presented in **Table 2**. Both IMRT and IMPT plans provided appropriate target coverage of the high-dose CTV (median V95 99.91% for both) and low-dose CTV (median V95 99.71% and 99.90% respectively). The IMPT plans had significantly lower maximum doses for both spinal cord and brainstem. The median maximum dose to the spinal cord was 37.3Gy in IMRT and 6.4Gy in IMPT (p<0.001), while median maximum dose to the brainstem was 33.0Gy in IMRT and 5.6Gy in IMPT (p<0.001). Mean doses to the contralateral parotid glands (median 7.6Gy vs 0.04Gy, p<0.001), contralateral submandibular glands (median 15.4Gy vs 1.4Gy, p<0.001), and contralateral pharyngeal constrictors (median 26.2Gy vs 20.4Gy, p<0.001) were significantly reduced with IMPT compared to IMRT. Furthermore, mean dose to the larynx, oral cavity, and lips were also significantly reduced with IMPT. Mean dose to the ipsilateral submandibular gland was minimally increased with IMPT compared to IMRT (median 62.5Gy to 61.6Gy, p<0.001). Of note, the maximum skin dose was not increased in IMPT compared to IMRT (median 66.1Gy vs 64.4Gy, p = 0.15).

**Table 2.** Target coverage and OAR doses.

| Structures | Median (range) |  | p-value |
| --- | --- | --- | --- |
|  | IMRT plan | IMPT plan |  |
| **Coverage, %** |  |  |  |
| High-dose CTV V95 | 99.91<br>(95.52-100) | 99.91<br>(98.19-100) | 0.72 |
| Low-dose CTV V95 | 99.71<br>(96.20-100) | 99.90<br>(99.56-99.99) | 0.022 |
| **Maximum dose, Gy** |  |  |  |
| Spinal cord | 37.3<br>(25.5-39.9) | 6.4<br>(4.8-18.8) | <0.001* |
| Brainstem | 33.0<br>(4.5-49.2) | 5.6<br>(0.7-24.6) | <0.001* |
| Skin rind | 64.4<br>(44.2-74.6) | 66.1<br>(47.3-80.1) | 0.15 |
| **Mean dose, Gy** |  |  |  |
| Esophagus | 12.4<br>(0.2-28.8) | 5.4<br>(0.008-36.2) | 0.004 |
| Larynx | 25.2<br>(0.8-43.1) | 20.8<br>(0.2-43.1) | 0.002* |
| Oral Cavity | 33.6<br>(17.8-64.0) | 19.7<br>(2.5-64.2) | <0.001* |
| Lips | 13.8<br>(7.3-36.8) | 1.7<br>(0.4-27.5) | <0.001* |
| Pharyngeal constrictor (ipsilateral) | 51.9<br>(19.7-68.9) | 50.2<br>(24.6-69.3) | 0.009 |
| Pharyngeal constrictor<br>(contralateral) | 26.2<br>(9.1-61.8) | 20.4<br>(3.9-58.1) | <0.001* |
| Parotid (ipsilateral) | 42.6<br>(12.9-71.4) | 44.2<br>(6.2-72.1) | 0.47 |
| Parotid (contralateral) | 7.6<br>(1.2-15.0) | 0.04<br>(0.00-3.0) | <0.001* |
| Submandibular gland (ipsilateral) | 61.6<br>(7.1-72.6) | 62.5<br>(17.8-73.1) | 0.001* |
| Submandibular gland<br>(contralateral) | 15.4<br>(2.5-51.1) | 1.4<br>(0.02-51.1) | <0.001* |

\*Bonferroni adjusted p-value = 0.005

IMRT and simulated IMPT plans for a man in his 50’s with p16+ T2N2bM0 oropharyngeal disease treated with post-operative IMRT are shown in **Figure 1**. A dose of 60Gy in 30 fractions was delivered to the high-risk CTV, and dose painting was used in the elective ipsilateral neck levels IB, II, III, IV and V to 52Gy. The IMPT colorwash dose distribution (**Figure 1C-D)** demonstrated sparing of the contralateral parotid from low-dose spill seen in the IMRT plan (**Figure 1A-B**). The corresponding dose-volume histograms are shown in **Figure 2**, confirming equivalence of CTV dose in both plans and reductions in dose to spinal cord, brainstem, contralateral parotid, and contralateral submandibular glands in the IMPT plan compared to the IMRT plan. For a second representative patient, IMRT/IMPT plans (**Figure 3**) and the corresponding dose-volume histogram (**Figure 4**) are displayed for a man in his 80’s with squamous cell carcinoma of the left post-auricular skin treated with post-operative IMRT to 66Gy and ipsilateral neck levels II, III, and IV to 54Gy. In this patient, the dose-volume histogram of the simulated IMPT plan highlighted reduced dose to the submandibular gland with IMPT.

**Figure 1.**
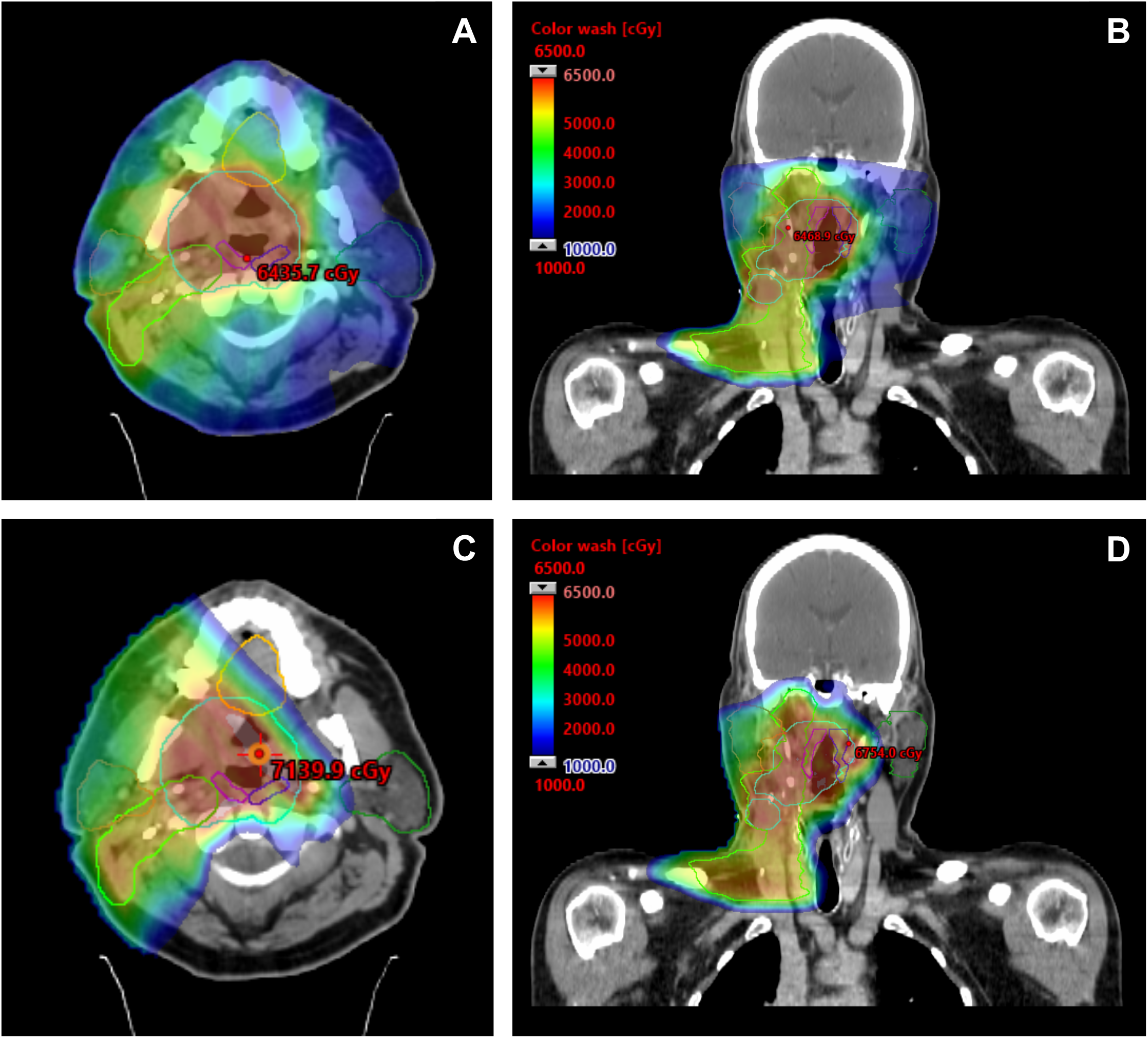
Representative color wash dose distribution for IMRT and IMPT plans. The patient had p16+ right-sided oropharyngeal disease and was treated with post-operative RT to 60 Gy in 30 Fx. Representative IMRT dose color wash in axial (A) and coronal (B) views. The corresponding simulated IMPT plan in axial (C) and coronal (D) views.

**Figure 2.**
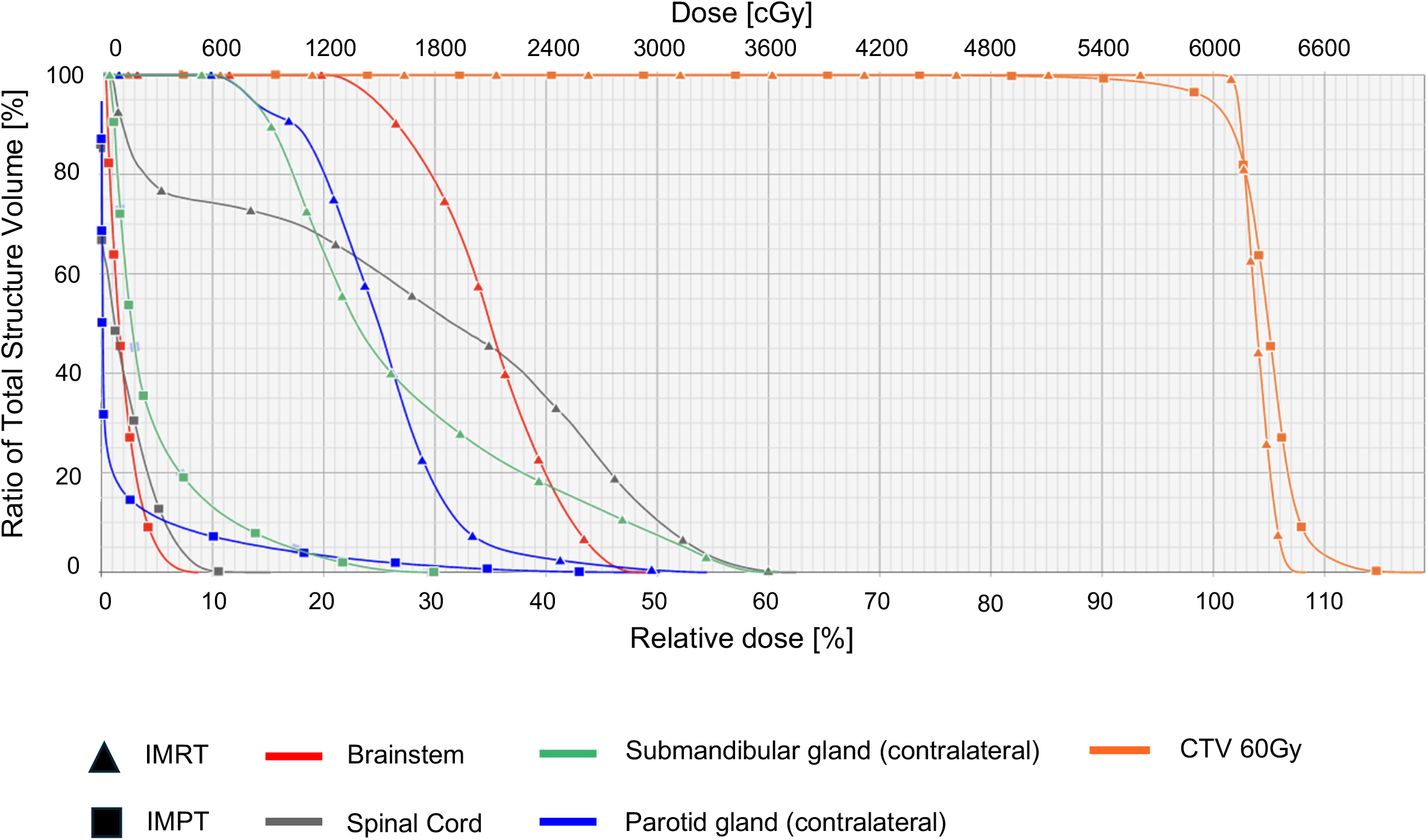
Dose-volume histogram in the same representative patient with oropharyngeal cancer. Dose received in the IMRT plan are represented with squares while doses from the simulated IMPT plan are represented with triangles.

**Figure 3.**
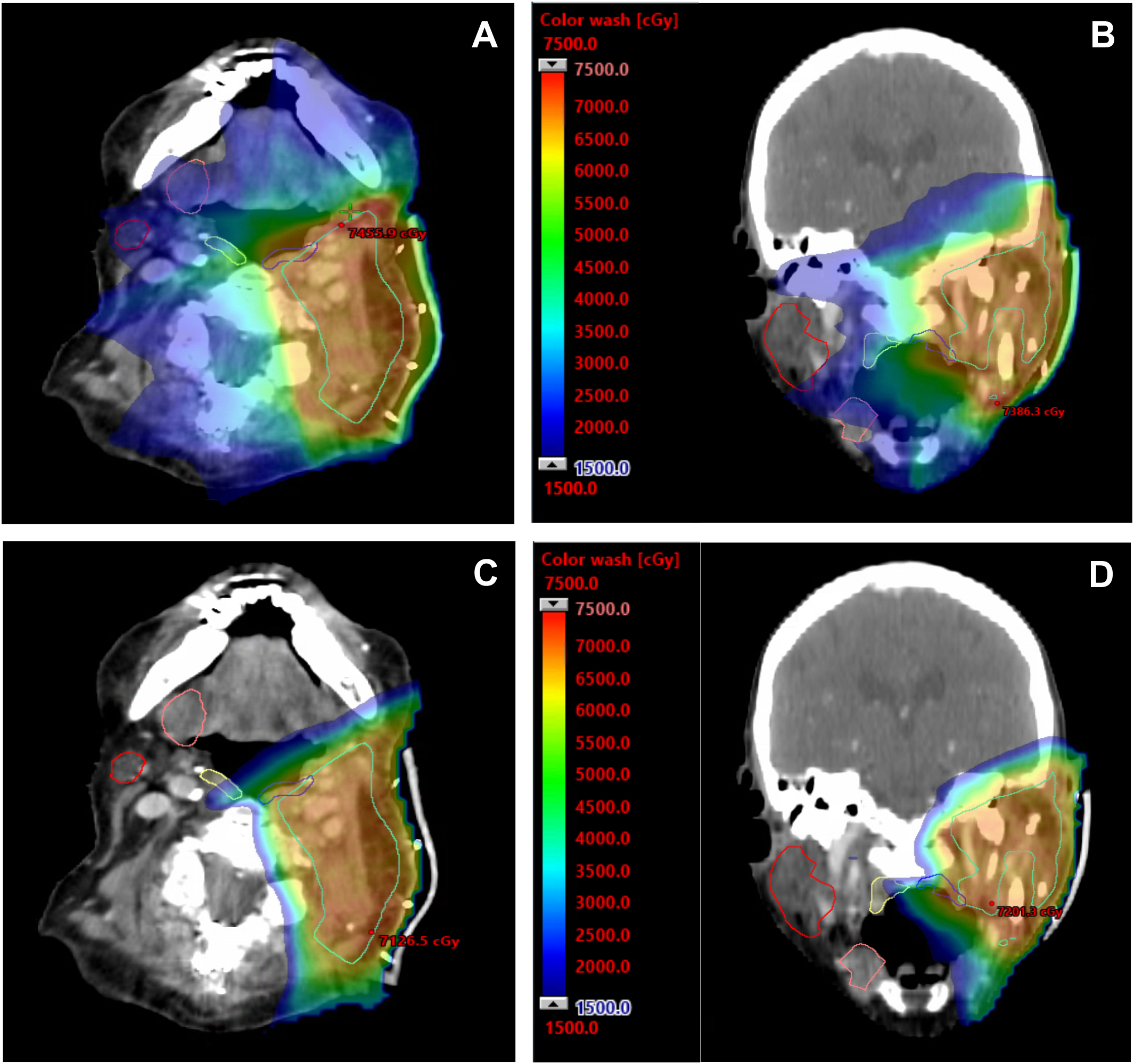
Representative color wash dose distribution for IMRT and IMPT plans in a second patient. The patient had squamous cell carcinoma of the skin and was treated with adjuvant RT to 66 Gy in 33 Fx. Representative IMRT dose color wash in axial (A) and coronal (B) views. The corresponding simulated IMPT plan in axial (C) and coronal (D) views.

**Figure 4.**
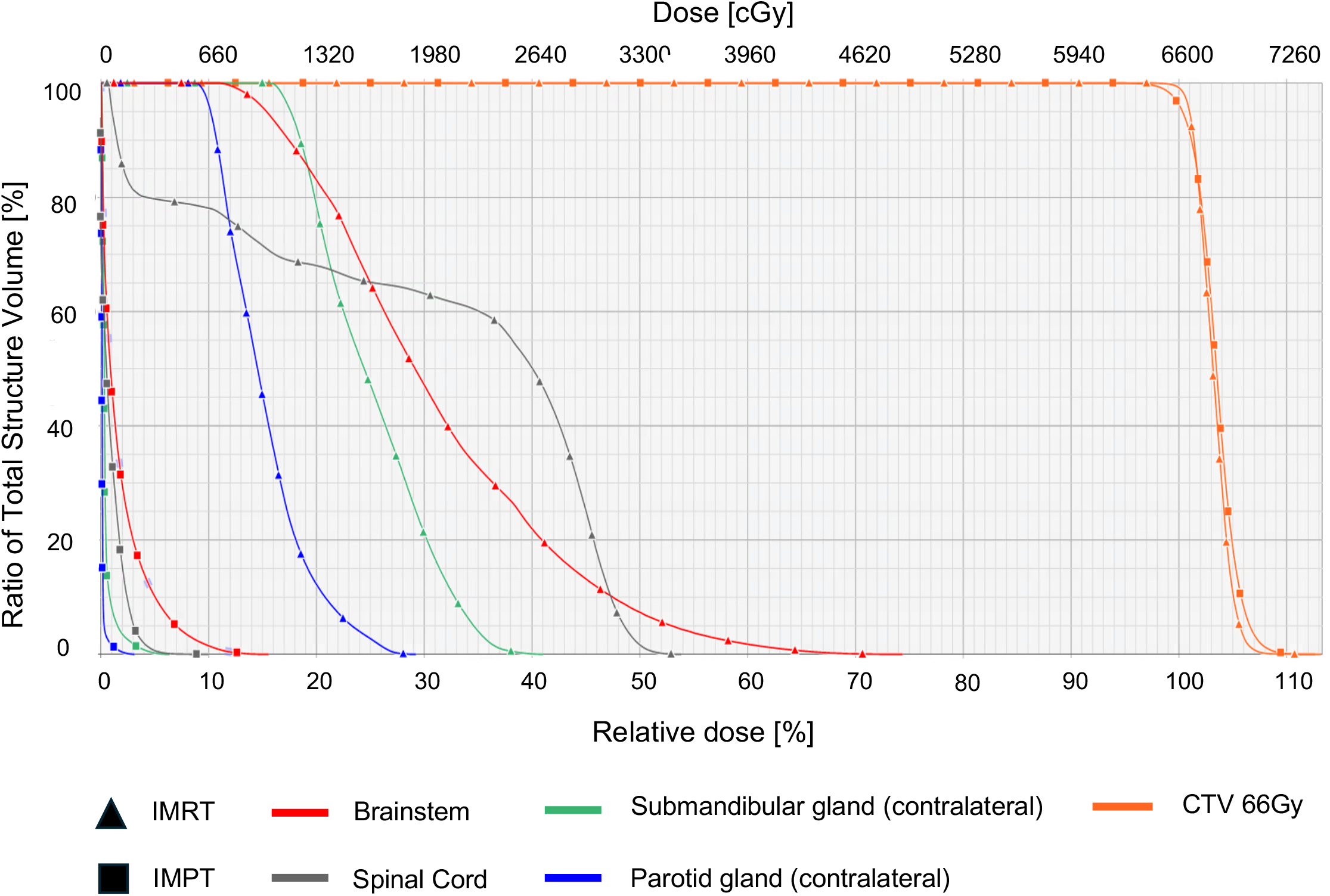
Dose-volume histogram in the same representative patient with squamous cell of the skin. Dose received in the IMRT plan are represented with squares while doses from the simulated IMPT plan are represented with triangles.

## DISCUSSION

In this dosimetric analysis among patients with deterioration of dysphagia symptoms after unilateral radiation for head and neck cancer, we examined differences in target coverage and OAR dose between delivered IMRT plans and simulated IMPT plans. By selecting patients who experienced significant toxicity after radiotherapy, we focused on a cohort that may particularly benefit from OAR sparing with IMPT. Our study demonstrated that IMPT provides appropriate target coverage while significantly reducing dose to critical structures, even among a cohort that predominately consisted of patients with locally advanced disease. IMPT plans reduced maximum dose to critical nervous system structures including the spinal cord and brainstem. Importantly, IMPT spared dose to contralateral OARs. Given that T3-T4 and/or N2-N3 disease often results in a greater dose to ipsilateral OARs (e.g., pharyngeal constrictors, submandibular gland, parotid gland) due to the extent of disease involvement, it becomes even more important to spare contralateral OARs to minimize toxicity for patients with locally advanced disease.

The majority of studies that have demonstrated a dosimetric benefit of IMPT in treating unilateral head and neck cancers have been limited to a single disease site. Our study corroborates the findings of other studies in multiple disease sites, including oropharynx, oral cavity, parotid gland, and skin. However, the clinical benefits of these dosimetric differences are largely unknown [6]. While mean dose to contralateral OARs, such as pharyngeal constrictors or major salivary glands, met all constraints with delivered IMRT plans by a significant margin, these patients nevertheless developed symptomatic toxicity. Multiple studies have proposed thresholds for plan optimization (e.g., mean parotid <26Gy), but the data also show linear dose-response extending below these thresholds, suggesting that dose should be kept as low as reasonably possible to reduce toxicity [19, 20].

Our study has limitations including its retrospective design in which IMPT plans were generated with *a priori* knowledge of the IMRT plans. Furthermore, there are limitations to proton therapy that can affect its clinical effect in practice. While protons are often not skin-sparing in the same way as photons are, our study did not demonstrate a significant difference in skin dose. Other considerations include greater range uncertainty compared to photon therapy, as well as a greater vulnerability to inter-fractional changes (e.g., weight change, tumor shrinkage) that can significantly alter dosimetry due to sharp distal dose falloff. Therefore, it is essential to prospectively compare IMRT and IMPT to assess the clinical impact of these two modalities. There is an ongoing phase II study comparing proton-vs photon-based post-operative radiation of unilateral salivary cancer, cutaneous squamous cell cancer, and melanoma at Memorial Sloan Kettering Cancer Center (NCT02923570). The various primary tumor sites included in our study supports similarly expanded inclusion criteria for other studies in the future.

## CONCLUSIONS

Among patients with majority locally advanced head and neck cancer receiving unilateral radiation, IMPT significantly decreases dose to critical central nervous system structures and contralateral OARs associated with acute and late toxicities. Future prospective studies will be critical to defining the role of IMPT in the treatment of unilateral head and neck cancer to reduce toxicities such as dysphagia and xerostomia.

## Data Availability

All data produced in the present study are available upon reasonable request to the authors.

